# Functional analysis of *LDLR* variants reveals that the novel p.(Ser610Tyr) variant in the YMDD motif drastically impacts LDL uptake

**DOI:** 10.1101/2025.05.16.25325576

**Authors:** Cris Ortiz López, Neila Rodríguez Roca, Irene Hidalgo Mayoral, Amanda Herranz Cecilia, Carlos Rodríguez Antolín, Pedro Martínez Hernández, Ana Carazo Álvarez, Francisco Jesús Arrieta Blanco, Sara Fernández Hernández, María Jesús Delgado-Martos, Carmen Rodriguez Jiménez, Sonia Rodríguez-Nóvoa

**Author notes:** **Corresponding authors:** Sonia Rodríguez-Nóvoa, Genetics of Metabolic Diseases Laboratory, Genetics Department, Hospital Universitario La Paz, Paseo de la Castellana 261, 28046, Madrid, Spain, Carmen Rodríguez Jiménez, Genetics of Metabolic Diseases Laboratory, Genetics Department, Hospital Universitario La Paz, Paseo de la Castellana 261, 28046, Madrid, Spain.

## Abstract

**Background and Aims:** Familial hypercholesterolemia (FH) is an inherited metabolic disease characterized by high low-density lipoprotein cholesterol (LDL-c) levels, leading to premature cardiovascular events. The heterozygous form (HeFH) affects 1 in 313 people, while the homozygous form (HoFH) affects 1 in 360,000. FH is primarily caused by pathogenic variants in the *LDLR* gene (79%), with other genes like *APOB, PCSK9*, and *LDLRAP1* also involved. Functional studies are essential to determine the impact of these variants on protein function, aiding in accurate diagnosis and treatment. This study characterizes a new *LDLR* variant and analyzes several others in hypercholesterolemia patients.

**Methods:** A newly detected variant, along with seven other *LDLR* variants initially classified as of uncertain significance, were selected from a cohort of hypercholesterolemic patients referred to our center for genetic analysis. Variant classification was performed according to ACMG/AMP guidelines. Functional studies included measuring LDL-Dil uptake, binding, and surface expression of the LDLR using flow cytometry.

**Results:** The analyzed variants included the novel, *LDLR*: c.1829C>A p.(Ser610Tyr) and the following: c.115T>C p.(Cys39Arg), c.185C>T p.(Thr62Met), c.968G>T p.(Gly323Val), c.1156G>T p.(Asp386Tyr), c.1966C>T p.(His656Tyr), c.2101G>A p.(Gly701Ser), and c.2231G>A p.(Arg744Gln). Functional studies reclassified five of these variants as probably pathogenic or pathogenic.

**Conclusions:** Functional studies of *LDLR* variants are essential to demonstrate disease causality in FH. Early diagnosis and appropriate treatment can significantly reduce coronary artery disease, highlighting the importance of demonstrating the impact of *LDLR* variants.

## Introduction

Familial hypercholesterolemia (FH; MIM# 143890) is an underdiagnosed inherited metabolic disease characterized by elevated low-density lipoprotein cholesterol (LDL-c) concentrations. FH significantly impacts global health due to its strong association with premature cardiovascular events. The heterozygous form (HeFH) has a prevalence 1 in 313 people in the general population, while the homozygous form (HoFH) occurs in 1 in 360,000 individuals [1] [2]. HeFH is characterized by LDL-c levels exceeding 190 mg/dL, whereas HoFH is associated with levels above ~400 mg/dL [3] [4].

The clinical phenotype of FH results from pathogenic variants in the following genes: *LDLR* (MIM# 606945) which codes for the LDL receptor (LDLR) and accounts for 79% of cases; *APOB* (MIM# 107730) coding for apolipoprotein-B (APOB) and responsible for 5% of cases; and *PCSK9* (MIM# 607786) coding for proprotein convertase subtilisin/kexine type 9 (PCSK9), which increases LDL receptor degradation and affects 1% of cases. All of these follow an autosomal dominant inheritance pattern. Additionally, the autosomal recessive form, caused by pathogenic variants in *LDLRAP1* (MIM# 605747), occurs in less than 1% of cases [5] [6].

Pathogenic variants in *APOE* disrupt lipoprotein metabolism pathways, influencing both cholesterol and triglyceride levels. Rare *APOE* variants have been linked to conditions such as lipoprotein glomerulopathy and familial combined hyperlipidemia (FCH), and homozygosity for the isoform E2 is associated with dysbetalipoproteinemia. Recent studies indicate that certain rare *APOE* variants contribute to both autosomal dominant hypercholesterolemia and FCH. Due to overlapping phenotypes and familial patterns, distinguishing FH from FCH can be challenging; fortunately, genetic testing aids in differential diagnosis [7].

The *STAP1* gene was initially proposed as a novel gene associated with FH; however, subsequent studies have questioned this role. More recently, its role as a causative gene for FH have been dismissed [8]. Variants in other genes, such as *ABCG5, ABCG8, CYP27A1* and *LIPA* are considered as FH phenocopies, providing additional information in genetic testing.

Currently, pathogenic variants in the *LDLR* gene are the primary cause of FH, accounting for over 90% of cases. The *LDLR* gene is located on chromosome 19 and comprises 18 exons. It encodes a protein with 860 amino acids. Exon 1 encodes an untranslated region with 21 hydrophobic amino acids that form the signal sequence [9]. This sequence is cleaved from the protein during its translocation into the endoplasmic reticulum (ER), resulting in a mature protein consisting of 839 amino acids [10]. LDLR is ubiquitously expressed and plays a crucial role in cholesterol homeostasis [11].

The LDLR is organized into functional subdomains, consisting of an ectodomain and an intracellular domain. The ectodomain includes the ligand-binding domain (LBD), the epidermal growth factor precursor homology domain (EGF), and the O-linked glycosylation domain. The intracellular domain contains the transmembrane and cytoplasmic regions. The LBD, encoded by exons 2 to 6, interacts with lipoproteins and corresponds to the amino terminal end of the protein. It consist of seven tandem repeats of 40-amino-acid (LR1 to LR7), each containing six conserved cysteine residues [12]. Each repeat is encoded by an exon, except for LR3, LR4, and LR5, which are all encoded by exon 4 [10]. Exons 7 to 14 encode a domain that shares 33% homology with the extracellular portion of the EGF domain. This domain includes two EGF-like repeats, EGF-A and EGF-B, separated by a β-propeller domain from a third EGF-like repeat, EGF-C [13] [14]. The EGF-A domain is crucial for PCSK9 binding at both neutral pH conditions on the cell surface and acidic pH conditions in the endosome [15]. The EGF-A domain alone drives PCSK9-mediated LDLR degradation, indicating its role in high-affinity interaction within the acidic endosomal environment. Additionally, the EGF domain is also involved in positioning the ligand-binding domain correctly on the cell surface [14]. Exon 5 encodes the O-linked glycosylation domain, located just above the transmembrane domain, with a sequence rich in Ser and Thr amino acids, which are abundant in O-glycosylations. Exon 16 and the final portion of exon 17 encode the transmembrane domain. The remaining part of exon 17 and exon 18 encode the cytosolic domain, which is involved in the endocytosis of the protein. This domain contains the NPXY sequence, important for receptor localization on the cell surface [9], and interacts with the phosphotyrosine-binding domain of the clathrin adaptor protein ARH, encoded by the *LDLRAP1* gene [5].

Genetic variants in *LDLR* can disrupt various stages of the LDLR-LDL endocytosis cycle, leading to FH. These variants can be classified accordingly with their impact [16] [17] [18]: class 1, includes variants that result in a nonfunctional or absent LDLR receptor; class 2, refers to variants that cause improper folding, leading to defective transport to the Golgi or plasma membrane, with retention in the endoplasmic reticulum (ER); class 3 involves variants that impair binding to ApoB; class 4, consists of variants that affect the internalization of LDLR, preventing its recruitment into clathrin-coated pits; class 5 describes variants causing recycling defects, which result in LDLR degradation in the lysosome due to improper LDL release in endosomes; class 6, includes variants that fail to transport LDLR to the basolateral membrane or prevent proper membrane insertion, leading to receptor secretion instead of its localization on the cell membrane.

More than 3400 distinct variants have been identified throughout LDLR in the ClinVar Database (ClinVar Miner, updated to 05/03/2025), including pathogenic, likely pathogenic and variants of unknown significance. Traditionally, the pathogenicity assessment of variants has been based on the standardized consensus guidelines developed by the American College of Medical Genetics and Genomics (ACMG) and the Association for Molecular Pathology (AMP) in 2015. According to their criteria, variants are classified as pathogenic, likely pathogenic, variant of unknown significance (VUS), likely benign and benign. To enhance precision and uniformity in variant interpretation, the Clinical Genome Resource Familial Hypercholesterolemia Variant Curation Expert Panel was assigned to refine and adapt the ACMG/AMP framework specifically for the classification of variants associated with FH [19]. Up to date, a large amount of variants remains as VUS or there is controversy regarding their classification. In this context, functional studies (PS3 criteria) aiming to assess their impact on the protein function are an indispensable resource of information for reclassifying variants of uncertain significance as pathogenic or benign. However, few functional studies are performed in newly identified variants, probably due to their labor-intensive nature.

In this study, we report and characterize a new genetic variant in the *LDLR* gene and conduct functional analysis of several *LDLR* variants identified in patients with hypercholesterolemia who were referred to our hospital for molecular testing.

## Material and Methods

### Selection of LDLR variants

A total of 8 *LDLR* variants were selected from a cohort of patients with hypercholesterolemia referred to our center for genetic analysis. Patients met the criteria of possible, probable or certain familial hypercholesterolemia according to the criteria of the Dutch Lipid Clinic Network [20]. All patients provided informed consent for genetic testing according to the routine testing protocols.

### Genetic analysis

Genomic DNA from probands was extracted from EDTA-treated whole blood samples using the Chemagen system (Chemagic DNA extraction special, Perkin Elmer Inc., Baesweiler, Germany). DNA quantification was carried out with a NanoDrop ND-1000 Spectrophotometer. Genetic analysis was performed using Next Generation Sequencing (NGS) with a custom panel of 198 genes. Library preparation and exome enrichment were conducted according to the manufacturer’s protocol (Nimblegen, Roche), followed by sequencing on the MiSeq or NextSeq platforms (Illumina). A subset of genes was selected: *LDLR, APOB, PCSK9, LDLRAP1, STAP1, ABCG5, ABCG8, APOE, LIPA* and *CYP27A1*. NGS data met the quality standards of our laboratory, with more than 30x coverage for 99% of the target regions.

All identified variants in FH related genes were classified according to ACMG guidelines. Those variants at the *LDLR* that showed controversy in their classification and those classified as variants of uncertain significant were included in subsequent functional studies. The class 2 variant p.(Gly549Asp) was used as a control, this variant has been functionally tested in fibroblasts from a homozygous patient, showing the receptor activity <2% [21]. Those variants not listed in the ClinVar, LOVD, or Human Gene Mutation Database (HGMD) were considered novel.

### In silico analysis

Bioinformatic analysis was conducted using algorithms developed by our bioinformatics unit. In summary, sequences were aligned to the CRCh37/hg19 human reference genome. Databases utilized for variant analysis included HGMD® (http://www.hgmd.cf.ac.uk/ac/index.php) from BIOBASE Corporation, Online Mendelian Inheritance in Man (OMIM, www.omim.org), and Gene Tests (www.genetests.org). Variant annotation was performed using Ensembl’s Variant Effect Predictor Tool.

In silico pathogenicity predictors employed were Combined Annotation Dependent Depletion (CADD), REVEL, Polymorphism Phenotyping (PolyPhen), MutAssessor, Fasthmm, and Vest. Conservation scores used included Gerp2, PhasCons, and Phylop across 13 species. For splicing prediction, tools such as MaxEntScan, NNSplice, GeneSplicer, and Human Splicing Finder were applied. The analysis files were uploaded in BAM format and displayed using Alamut Visual V.2.8.0 (Interactive Biosoftware, France).

### Cell culture and transfection

Chinese hamster ovary (CHO) *ldl*A7 cells (kindly provided by Dr. Monty Krieger, Massachusetts Institute of Technology, Cambridge, MA) were used as the *LDLR*-deficient cell model [22] [23]. These cells were cultured in Ham’s F-12 medium (Gibco, Life Technologies, Carlsbad, CA) supplemented with 5% heat-inactivated fetal bovine serum (Gibco, Life Technologies), 100 U/ml penicillin, 100 µg/ml streptomycin, 2 mmol/L L-glutamine, and 50 mg/ml Normocin (InvivoGen, Toulouse, France). The CHO-*ldl*A7 cells were grown as adherent cultures in 75 cm^2^ flasks and incubated at 37°C in a 5% CO_2_ atmosphere.

The constructs were created with the LDLR NM_000527 human cDNA ORF clone expression vector with a C-GFPSpark® tag (HG10231-ACG Sino-biological, Wayne). Site-directed mutagenesis was performed using the QuikChange site-directed mutagenesis lightning kit (Agilent Technologies) as per the manufacturer’s protocol. Sanger sequencing confirmed the plasmid sequences following mutagenesis.

CHO-*ldl*A7 cells were plated in 24-well plates at a density of 15 × 10□ cells per well, and once they reached 70-90% confluency, they were transiently transfected. Lipofectamine 3000 (Invitrogen™ Thermo Fisher Scientific, Waltham) was used for transfection, with 2.5 µg of each *LDLR* expression plasmid added. Cells were also transfected with an empty vector and wild type (WT) plasmid, following the manufacturer’s instructions. The transfection mixture was applied to the cells, and incubation was carried out for 48 hours.

### Quantification of LDL-receptor activity by flow cytometry. Uptake and binding of LDL-Dil

Human LDL was isolated from a single donor and labeled with 1,1′-dioctadecyl-3,3,3,3′-tetramethylindocarbocyanine perchlorate (DiI; Molecular Probes, Life Technologies; Calvo, Gómez-Coronado, Suarez, Lasuncion, & Vega, 1998). Transfected cells were incubated for 4 hours at 4°C to assess binding, and at 37°C to evaluate uptake, using 20 µg/ml of LDL-Dil protein. Following incubation, cells were washed twice with phosphate-buffered saline containing 2% bovine serum albumin (PBS-2% BSA) and analyzed via flow cytometry (CytExpert Cytoflex™). Forward and side scatter gates were set to exclude dead cells and debris, and 4′,6-diamidino-2-phenylindole dihydrochloride (DAPI; Invitrogen™, Thermo Fisher Scientific) was added at a final concentration of 0.1% to differentiate live from dead cells. A total of 10□ cells were acquired for each analysis. All experiments were conducted in triplicate.

### Quantification of LDLR expression by flow cytometry

After 48 hours of transfection, the cells were labeled with anti-hLDLR (APC: R&D Systems, Novus Biologicals, CO, USA) at a concentration of 0.3125 µg/ml in PBS-1% BSA for 20 minutes at room temperature and in darkness. After this incubation, the labeling process was stopped by adding 2 ml of PBS. The cell pellet was suspended in PBS, kept on ice, and in darkness until analysis by the CytExpert Cytoflex™ flow cytometer. Forward and side scatter gates were set to exclude dead cells and debris, and 4′,6-diamidino-2-phenylindole dihydrochloride (DAPI; Invitrogen™, Thermo Fisher Scientific) was added at a final concentration of 0.1% to differentiate live from dead cells. A total of 10L cells were acquired for each analysis. All experiments were conducted in triplicate.

## Results

All *LDLR* variants tested were identified in index patients. We discovered a new variant in *LDLR*, ***c*.*1829C>A*; p.(Ser610Tyr**), located in exon 12. Additionally, seven other *LDLR* variants with controversial classifications were identified and selected for functional analysis: *c*.*115T>C*; p.(Cys39Arg), *c*.*185C>T*; p.(Thr62Met), *c*.*968G>T*; p.(Gly323Val), *c*.*1156G>T*; p.(Asp386Tyr), *c*.*1966C>T*; p.(His656Tyr), *c*.*2101G>A*; p.(Gly701Ser), and *c*.*2231G>A*; p.(Arg744Gln). **Figure 1** shows the locations of these *LDLR* variants, and the characteristics of the variants and the index patients are detailed in **Table 1**.

**Table 1.** Characteristics of patients and classification of variants found in *LDLR* according FH VCEP guidelines (ACMG– American College of Medical Genetics) [26]. Abbreviations: CVD: cardiovascular disease; FS: functional study; ID: proband identification; LDL-c: low-density lipoprotein cholesterol; nd: no data available; VUS: variant of unknown significance. *Note*. New variants at *LDLR* are marked in bold.

| ID | Sex | Family history |  | Personal history | Physical examination |  | LDL (mg/dL) | Exon | cDNA | Protein | LDLR domain | Reference | ACMG Classification prior FS | Final ACMG Classification |
| --- | --- | --- | --- | --- | --- | --- | --- | --- | --- | --- | --- | --- | --- | --- |
|  |  | Increased LDL-c | ECV | ECV | Xanthomas | Arcus cornealis |  |  |  |  |  |  |  |  |
| 1 | M | nd | nd | nd | nd | nd | >180 | 2 | c.115T>C | p.(Cys39Arg) | Ligand binding | [31] | VUS | **Likely Pathogenic** (PS3, PM1, PM2, PP3) |
| 2 | F | Yes | Yes | No | No | No | 250-329 | 2 | c.185C>T | p.(Thr62Met) | Ligand binding | [32] | VUS | **VUS** (PP1, PS3_supporting) |
| 3 | F | No | Yes | No | Yes | No | 190-249 | 7 | c.968G>T | p.(Gly323Val) | EGF precursor homology | [38] | VUS | **Pathogenic** (PS3, PM2, PP1, PP3, PP4, PS4_supporting) |
| 4 | M | nd | Yes | nd | nd | nd | 250-329 | 8 | c.1156G>T | p.(Asp386Tyr) | EGF precursor homology | [39] | VUS | **Pathogenic** (PS3, PM2, PS4_moderate, PP3, PP4) |
| 5 | F | Yes | No | No | No | No | 250-329 | 12 | c.1829C>A | **p.(Ser610Tyr)** | EGF precursor homology | **New** | VUS | **Likely Pathogenic** (PS3, PM2, PP3, PP4) |
| 6 | M | Yes | Yes | No | No | No | 250-329 | 13 | c.1966C>T | p.(His656Tyr) | EGF precursor homology | [41] | VUS | **Likely Pathogenic** (PS3, PM2, PP3, PP4) |
| 7 | M | Yes | Yes | No | No | No | 250-329 | 14 | c.2101G>A | p.(Gly701Ser) | EGF precursor homology | [32] | VUS | **VUS** (PP1_Moderate, PP3) |
| 8 | F | nd | nd | nd | nd | nd | >180 | 15 | c.2231G>A | p.(Arg744Gln) | O-linked sugar | [41] | VUS | **Likely benign** (PS3_supporting, BP2, BP4) |

**Figure 1.**
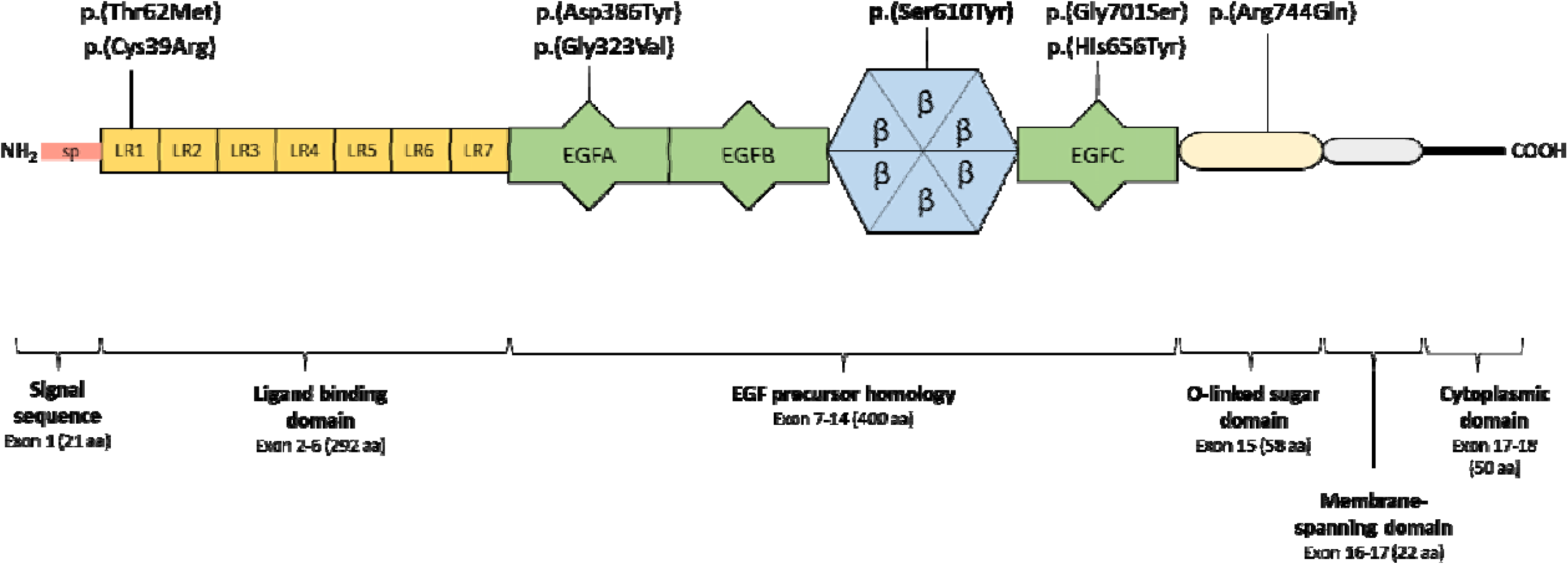
Location of the *LDLR* variants identified in this study. The new variant is highlighted in bold. Abbreviations: aa, amino acid; EGF, epidermal growth factor; LR, ligand-binding repeat; sp, signal peptide.

### Effect of the variants on the LDLR function

Effect of the LDLR variants on LDLR function was assessed in transfected cells with the different constructs by measuring LDL-Dil uptake and binding capacity through flow cytometry. The specific median fluorescence intensity for each sample was calculated by subtracting the fluorescence intensity of the empty vector, and the results were expressed as a percentage relative to the wild type (WT). The variants p.(Gly323Val) and p.(Ser610Tyr) showed less than 10% of LDL-Dil binding and uptake compared to the WT. The other variants had the following uptake and binding percentages, respectively: p.(Cys39Arg) 41% and 25%, p.(Thr62Met) 71% and 87%, p.(Asp386Tyr) 62% and 80%, and p.(His656Tyr) 61% and 100% (Figure 2). In contrast, the variants p.(Gly701Ser) and p.(Arg744Gln) showed 96% and 85% uptake, respectively, with binding efficiencies of 89% and 80%, which are closely comparable to the WT (**Figure 2**).

**Figure 2.**
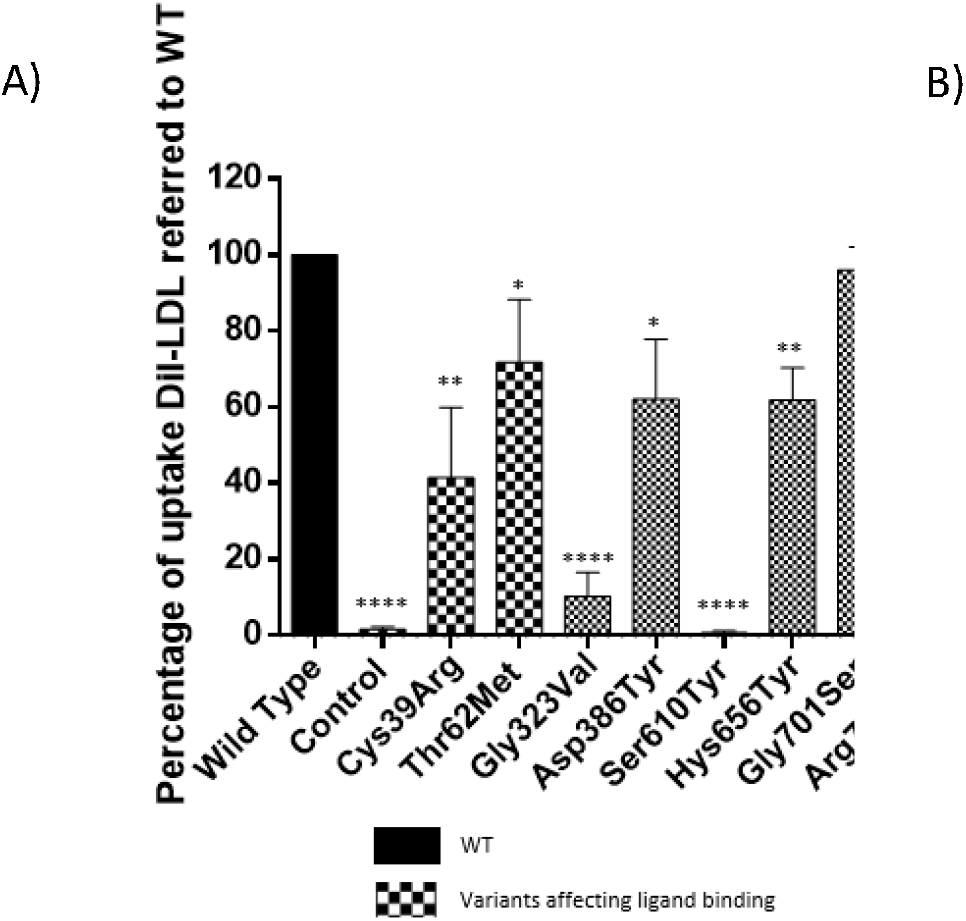
LDLR activity in the transiently transfected CHO-*ldl*A7 cells, with WT, control (Gly549Asp), and *LDLR* variants. A) Uptake LDL-Dil; B) Binding LDL-Dil. The analysis performed by flow cytometry as described in Material and Methods. Each result corresponds to mean□±□S.E.M. of the triplicate experiments compared with the WT. Statistical significance were determined by a two-tailed Student’s t test and a 95% confidence interval. LDL, low-density lipoprotein; WT, wild type. *p□<□.05, **p□<□.01, ***p□<□.001, ****p□<□.0001 versus WT. The results were obtained in three independent experiments.

### Surface expression of LDLR

Significant differences in LDLR expression compared to the WT were observed for two variants: p.(Cys39Arg) and p.(Ser610Tyr), with expression levels of 34% and <2%, respectively. The other variants, p.(Thr62Met), p.(Gly323Val), p.(Asp386Tyr), p.(His656Tyr), p.(Gly701Ser), and p.(Arg744Gln) showed surface LDLR expression levels relative to the WT of 93%, 44%, 80%, 100%, 86%, and 84%, respectively (**Figure 3**).

**Figure 3.**
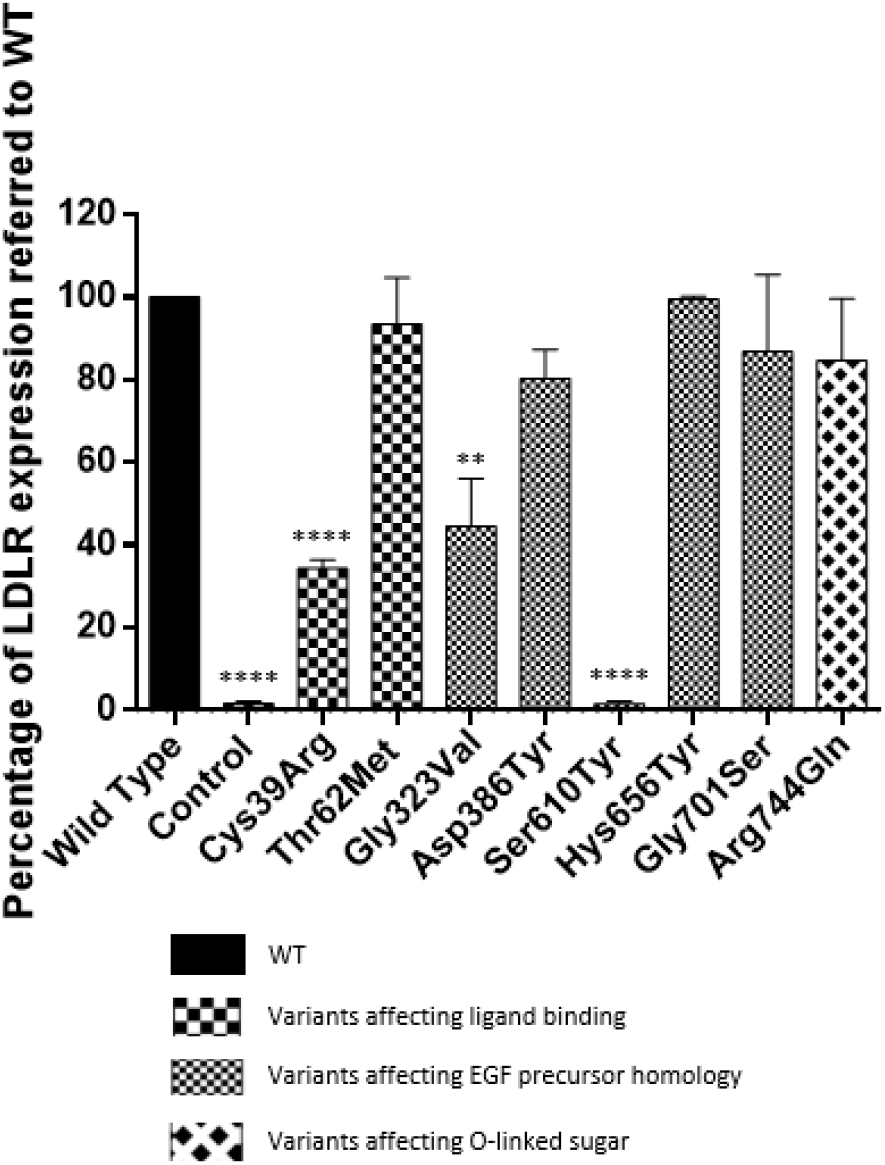
Surface expression of *LDLR* in transiently transfected CHO-*ldl*A7 cells with WT, control (Gly549Asp), and *LDLR* variants. Each result represents the mean ± S.E.M. of experiments conducted in triplicate compared to WT. Statistical significance was determined using a two-tailed Student’s t-test with a 95% confidence interval. *p < 0.05, **p < 0.01, ***p < 0.001, ****p < 0.0001 versus WT. The results were obtained from three independent experiments.

### Cosegregation studies

Cosegregation studies were conducted in three index cases: ID2, ID3, and ID4. The proband ID2 carried the variant p.(Thr62Met) and had LDL-c levels ranging from 250 to 329 mg/dL. Her father was not tested biochemically or genetically; however, her 51-55 years-old sister, diagnosed with hypercholesterolemia and undergoing lipid-lowering treatment, was also a carrier of the same variant. Her mother was hypercholesterolemic, and her brothers were normocholesterolemic, but none of the three were genetically tested (Figure 4A). The variant appeared to exhibit cosegregation with the disease phenotype; however, definitive evidence could not be established.

**Figure 4.**
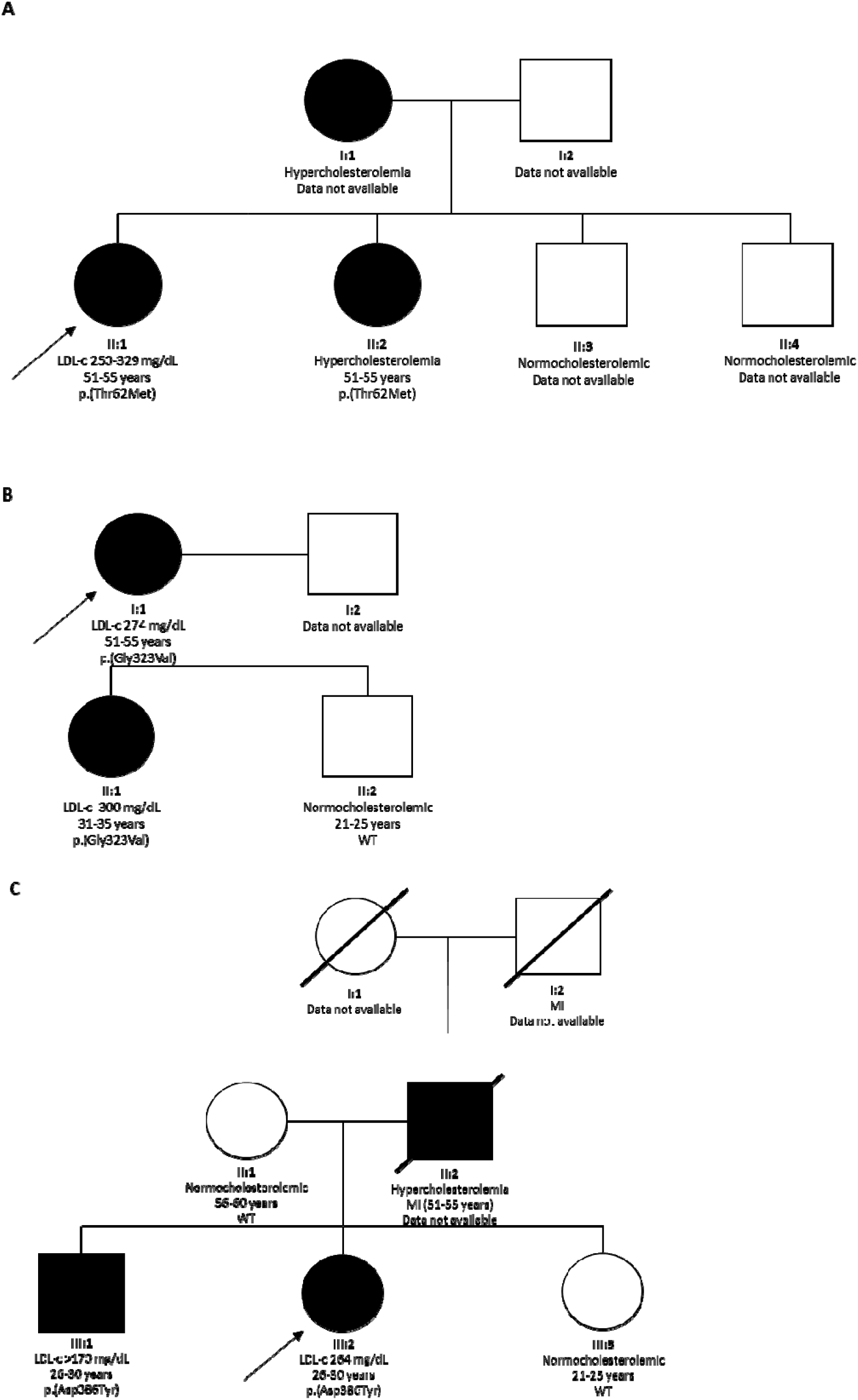
Pedigrees of families with validated missense variants in the *LDLR* gene. Index cases are indicated by an arrow, circles represent females and squares represent males. Symbols filled in black denote affected patients. The first line beneath the symbols corresponds to the family generation, while the second line presents the baseline LDL-c levels when these data are available. The third line indicates the age range, and the fourth line indicates the *LDLR* genotype or wild type (WT). **A**: Relatives of patient ID2 p.(Thr62Met). **B**: Relatives of patient ID3 p.(Gly323Val). **C**: Relatives of patient ID4 p.(Asp386Tyr).

The proband ID3 carried the variant p.(Gly323Val), with LDL-c levels of 274 mg/dL. Her daughter, who showed hypercholesterolemia, was also a carrier of the familial variant, while her 21-25 years-old son, who had a normocholesterolemic profile, did not carry the variant. The family pedigree showed cosegregation with the disease (Figure 4B), which was further supported by significant differences in binding and internalization observed in functional assays.

Lastly, the proband ID4 carried the variant p.(Asp386Tyr). In this family, both the paternal grandfather and father passed away due to acute myocardial infarction (MI). The genetic study was perfomed in the patient’s mother and two siblings. Both the father and brother exhibited elevated LDL-c levels, but only the brother was confirmed as a carrier of the variant, as the father was not gentically tested. In contrast, the mother and daughter, who had normocholesterolemic profiles, did not carry the variant (Figure 4C). While the variant appeared to cosegregate with the disease phenotype, this relationship could not be conclusively confirmed.

## Discussion

Numerous genetic variants in the *LDLR* gene have been identified as contributors to familial hypercholesterolemia. In this study, we characterized one novel variant alongside seven previously reported variants, whose impacts remain controversial.

In this study, we characterized two variants in LR1 module, **p.(Cys39Arg)** and **p.(Thr62Met)**. LR1 is part of the ligand-binding domain and initial studies have suggested that LR1 does not play a significant role in lipoprotein binding. However, subsequent research has shown that lipoprotein binding is likely facilitated by the collective action of multiple LRs, including LR1 [24]. While complete deletion of LR1 does not affect lipoprotein binding, specific missense variants within LR1 can significantly impact this process [25]. Notably, the number of variants identified in LR1 is lower compared to those in LR2-LR7. The **p.(Cys39Arg)** was previously described in a cohort of patients from Nevada [26] and lacked functional studies. This variant affects the Cys39, the third cystein of the LR1, preventing the formation of the disulfide bridge. As a result, the protein may not fold properly, leading to inadequate binding to ApoB. Functional analysis in our study showed a marked reduction in LDL-Dil binding, internalization, and surface expression of LDLR. The second variant, **p.(Thr62Met)**, was first reported in a cohort of patients with autosomal dominant hypercholesterolemia from the Netherlands [27]. The functional impact of this variant has been controversial, with studies ranging from benign to pathogenic. Two initial functional studies evaluated the impact of the p.(Thr62Met) variant. The first, conducted in 2015 by Thormaehlen et al. [28], deemed the variant non-deleterious; however, this approach was later dismissed by guidelines for not using heterologous cellular models to test LDLR functionality. The second, by Benito-Vicente et al. in 2018 [29], assessed LDLR expression and activity in CHO-*ldl*A7 transfected cells, using two methodologies to confirm LDLR expression and functionality comparable to the WT. In 2022, Graça et al. [30] included the p.(Thr62Met) variant as a control, with binding assays showing results between 70% and 90%, which were considered inconclusive. Since p.(Thr62Met) does not affect cysteines residues or the SDE sequence critical for calcium binding [31], it is not expected to impact binding. Our functional analysis demonstrated minimal effect on uptake, binding and LDLR expression compared to the WT.

The **p.(Gly323Val)** and **p.(Asp386Tyr)** variants are located at the module A of the EGF precursor homology domain [32], a highly conserved domain where variants could affect receptor function. Functional analysis of **p.(Gly323Val)** revealed a marked decrease in LDLR activity and expression. This variant was previously classified as variant of uncertain significance [33], but based on our study it can now be reclassified as likely pathogenic. The **p.(Asp386Tyr)** variant, which was previously described [34], also showed reduced activity in our analysis. This variant resulted in a 62% decrease in LDL uptake, leading to its reclassification from VUS to likely pathogenic. Both variants impair LDLR functionality, likely due to their impact on the EGFA domain, which plays a critical role in PCSK9-mediated degradation. Several variants in the EGFA domain have been described that increase PCSK9 binding, thereby reducing LDLR. Additionally, given the EGFA domain’
ss role in receptor positioning, the p.(Asp386Tyr) variant could alter protein folding during internalization, further compromising receptor-mediated LDL uptake.

The **p.(Ser610Tyr)** variant affects the β-propeller domain, which is involved in receptor recycling and lipoproteins release at low pH. The β-propeller domain consists of six YWTD repeats, which fold into a β-helix structure that controls various processes, including lipoprotein uptake and extracellular signal transduction [32]. Because this variant affects a critical region of the receptor, it is predicted to severely impact protein folding, potentially preventing the receptor from reaching the cell surface. Our study showed that p.(Ser610Tyr) resulted in null LDLR activity, with both uptake and binding, as well as surface expression, below 2%. This finding is consistent with the very high LDL-c levels observed in the patient (250 to 329 mg/dL). Other variants afecting the same aminoacid Ser610 have been reported in Clinvar associated with hypercholesterolemia, including p.(Ser610Pro) and p.(Ser610Cys), both classified as likely pathogenic, and p.(Ser610Phe) reported as pathogenic/likely pathogenic. These findings highlight the importance of this domain. Functional validation has been performed for other variants affecting this domain, yielding results similar to those observed in this study. For example, previous research of our group showed the p.(Tyr532Asn) variant had less than 1% of LDL-Dil binding and uptake [35]. Based on these findings, the p.(Ser610Tyr) variant is classified as likely pathogenic according to ACMG guidelines.

We also analyzed two variants located in the EGFC domain, a highly conserved region. Since the variants are not located in the linker between the β-helix and the EGFC, they are unlikely to affect the receptor folding and function. However, it has been suggested that the integrity of the interface between the helix and the EGFC module can be modulated by a decrease in pH triggered by ligand release in the endosome [32]. For the **p.(His656Tyr)** variant, LDL uptake was significantly lower compared to WT, with no differences in LDLR expression or LDL-Dil binding. This variant has been reported in Norwegian patients and their relatives [36]. The **p.(Gly701Ser)** variant was predicted to be potentially pathogenic by in silico analysis; however, but cosegregation studies have yielded mixed results. Cosegregation was observed in two families, but another family did not exhibit cosegregation [37], highlighting the variability in findings. In our functional study, the variant showed minimal impact on LDL-Dil uptake, binding and LDLR expression.

The **p.(Arg744Gln)** variant, located in the O-linked sugar domain, was predicted to have a benign impact based on in silico analysis. It has been reported in FH populations, but no level 1 functional studies have been conducted [38]. This case is particularly perplexing as it fulfills contradictory ACMG criteria. On one hand, it exhibits pathogenicity indicators, such as being reported in at least 15 unrelated patients with FH and showing less than 85% LDLR expression in heterozygous patient lymphoblasts. On the other hand, it meets benign criteria, including its population frequency, its occurrence in a heterozygous FH patient carrying another pathogenic variant in the *APOB* gene, and evidence from a high-throughput microscopy assay indicating no disruption in expression and activity. Our assays results were consistent with these findings, showing LDLR activity and expression levels comparable to the WT.

In conclusion, we identified and characterized a novel *LDLR* variant located in the β-propeller domain that showed a strong deleterious effect. Additionally, four variants were reclassified from VUS to likely pathogenic or pathogenic, enhancing the effectiveness of molecular diagnosis of FH.

Functional studies of genetic variants are essential to establish disease causality in familial hypercholesterolemia. Understanding the impact of the genetic variants allows clinicians to make accurate diagnoses and tailor treatments to reduce coronary artery disease risk. Molecular testing’s diagnostic yield improves when variants are classified with sufficient evidence, reducing genetically unconfirmed cases. This study demonstrates that functional analysis enables the reclassification of *LDLR* variants, highlighting the importance of these studies in confirming genetic diagnoses, particulary given the high polymorphism of *LDLR*.

## Data Availability

All data produced in the present study are available upon reasonable request to the authors

